# Comparing Misophonia with Clinical Emotional Dysregulation: A Multi-method Characterization Study

**DOI:** 10.64898/2026.11.16.26368227

**Authors:** Z.E. Brasher, J.Y. Choi, S.C. Napolitano, A.D Neacsiu

## Abstract

**Background:** Misophonia, a condition characterized by intense distress in response to specific everyday sounds, is increasingly recognized as distinct from established auditory and psychiatric disorders. Although it is associated with emotional dysregulation, impairment, distress, and atypical reactivity relative to clinical controls, few studies have directly compared misophonia with a diverse clinical sample exhibiting general difficulties managing emotions. This limits the understanding of its underlying mechanisms and hinders the development of targeted interventions. To address this gap, we conducted a multimethod study comparing adults with moderate-to-severe misophonia and adults with clinically significant emotion dysregulation.

**Methods:** As part of a randomized controlled clinical trial, community-dwelling adults with elevated misophonia distress (n = 30) or clinically significant emotion dysregulation (n = 30) were enrolled in a study to examine sound responsiveness in misophonia. Participants completed a semi-structured clinical interview (SCID-5) and self-report measures assessing misophonia, emotional dysregulation, regulation, emotional processing, psychopathology, and contextual stress at intake. Next, physiological indices of arousal and regulation via skin conductance level (SCL), skin conductance response (SCR), and high-frequency heart rate variability (HF-HRV) were recorded during an Evocative Sound Challenge Task.

**Results:** At baseline, participants with misophonia reported lower emotion dysregulation than the emotion dysregulation group, despite comparable self-report impairment attributable to emotional difficulties. Physiologically, participants with misophonia exhibited higher baseline HF-HRV than the emotion dysregulation group (*p* = .010). There was no evidence of a baseline difference in SCL (*p* = .157) or SCR (*p* = .085). During and after the sound task, groups differed in HF-HRV (*p*s = .002-.017), with the clinical group exhibiting higher HF-HRV than the misophonia group. No group differences were observed in SCL or SCR during and after the sound task after correcting for baseline (all *p*s > .08).

**Conclusion:** Findings suggest that experiences of emotion dysregulation are relevant to misophonia but may differ from more generalized difficulties observed in clinically dysregulated populations. Despite endorsing substantial impairment, individuals with misophonia demonstrated lower global emotion dysregulation and greater emotion regulation self-efficacy, while physiological findings revealed distinct patterns of resting and task-related parasympathetic functioning. Collectively, the findings suggest that emotional difficulties in misophonia may be more context-dependent and closely tied to personally salient trigger cues than to generalized deficits in emotion regulation. Conceptualizing misophonia as pervasive emotion dysregulation may therefore conflate clinical impairment with the processes underlying symptom expression and obscure more targeted intervention mechanisms.

---

In misophonia, otherwise ordinary sounds acquire unusually strong emotional salience, eliciting intense physiological arousal and reactions such as anger, irritation, disgust, and urges to escape the sounds (Armstrong et al., 2023; Jager et al., 2020; Schröder et al., 2013; Swedo et al., 2022). These noises, often referred to as “trigger” sounds, are most commonly human-produced. Given the ubiquity of sound in daily activities, misophonia substantially impairs social and occupational functioning and reduces quality of life (Möllmann et al., 2023). While misophonia has often been conceptualized as a form of heightened sensory sensitivity (Apaydın et al., 2026; Woolley et al., 2025), misophonic responses do not appear to depend solely on the loudness or acoustic properties of sounds (Rasouli et al., 2026). Despite growing evidence suggesting that emotional, cognitive, and regulatory processes contribute to disorder presentation (Guetta et al., 2022; Kumar et al., 2017; Woolley et al., 2026), it remains unclear whether these findings reflect a broader vulnerability to emotion dysregulation or distress associated with a highly cue-specific condition. Consequently, it remains unclear how much emotion dysregulation associated with misophonia resembles broader clinical emotion dysregulation.

Multiple findings suggest that emotional dysfunction in misophonia may be relatively selective and context-dependent. Trigger responses are influenced by the sensory characteristics of sounds and accompanying visual cues (Rasouli et al., 2026), but also by contextual and interpersonal factors. Indeed, neurobiological and behavioral evidence suggests that triggers function as highly salient stimuli whose emotional significance depends on personal meaning and context (Kumar et al., 2017). For example, noises produced by family members are often experienced as more distressing than identical sounds generated by others (Edelstein et al., 2013), and distress may be reduced when the same sounds are paired with incongruent visual information (Samermit et al., 2022). Consistent with this interpretation, misophonic reactions are typically restricted to specific triggers and contexts rather than pervasive across situations (Edelstein et al., 2020) and many individuals with misophonia do not meet criteria for other psychiatric conditions (Swedo et al., 2022). Collectively, these findings suggest that distress in misophonia may be closely tied to personally salient triggers rather than reflecting a generalized pattern of emotional dysfunction.

At the same time, several lines of evidence suggest that emotional difficulties in misophonia may extend beyond immediate reactions to trigger sounds. Emerging evidence indicates that cognitive processes appear to be associated with both the persistence and generalization of distress associated with trigger sounds (Woolley et al., 2026). In a qualitative study, individuals with misophonia commonly report self-critical thoughts, negative appraisals of others, intrusive aggressive thoughts, and urges to escape during trigger exposure, followed by guilt, concerns about social evaluation, and repetitive recollections of prior triggers (Woolley et al., 2026). Consistent with these observations, misophonia has been associated with greater rumination and reduced cognitive flexibility (Black et al., 2026). Neurobiological findings further suggest hyperconnectivity across emotion, salience, and attentional networks, alongside evidence of reduced cognitive flexibility in adults with misophonia (Neacsiu et al., 2022).

In addition to the distressing cognitions extending beyond triggers, individuals with misophonia report elevated difficulties with emotion regulation (Dixon et al., 2024; Guetta et al., 2022), including difficulties with impulse control, emotional awareness, and access to effective regulation strategies (Erfanian et al., 2018). Supporting the clinical relevance of these deficits, impulsive behavior during distress partially mediates the relationship between neuroticism and misophonic distress (Cassiello-Robbins et al., 2020), while anxiety severity partially mediates the relationship between misophonic distress and reduced quality of life (Ay et al., 2024).

Additionally, misophonia has also been associated with elevated rates of anxiety, mood, and obsessive-compulsive related disorders (Freshley et al., 2026; Rosenthal et al., 2022), suggesting overlap with disorders characterized by broader emotion dysregulation. Further, internal states, such as stress, fatigue, physical discomfort, intrusive thoughts, and heightened emotional arousal have been reported to exacerbate misophonic trigger responses (Woolley et al., 2024). Likewise, emerging treatment research suggests that interventions that restructure cognitions around triggers may significantly reduce misophonic distress (Neacsiu et al., 2024). Collectively, these findings suggest that emotion regulation processes may contribute to the experience and maintenance of misophonic distress beyond the immediate reactions to trigger sounds.

Despite growing evidence linking emotion regulation difficulties to misophonia, it remains unclear whether these difficulties resemble the broader dysfunction observed in clinical populations. Emotion dysregulation is a transdiagnostic construct implicated across a range of psychiatric conditions and is characterized by difficulties in emotional awareness, modulation, and behavioral control (Gratz & Roemer, 2004; Gross, 1998). Given the magnitude of the anger, irritation, and disgust reactions documented in misophonia (Savard et al., 2022; Siepsiak et al., 2023) and the level of impairment observed from the condition, it is understandable that parallels might be made to broader emotion dysregulation and other psychiatric conditions. Still, existing research has largely relied on comparisons between individuals with misophonia and non-clinical control participants. Because non-clinical controls differ from individuals with clinically significant emotional dysfunction in many domains, such comparisons cannot determine whether heightened emotion dysregulation observed in misophonia is unique or reflects broader clinical distress. If emotional difficulties in misophonia reflect broader emotion dysregulation, individuals with misophonia would be expected to resemble clinically dysregulated populations across emotional and clinical domains. Alternatively, if emotional difficulties in misophonia are more context-dependent than uniformly generalized, meaningful differences between these groups would be expected to emerge. Uncertainty regarding the nature of emotion dysregulation in misophonia may complicate efforts to develop and evaluate targeted interventions.

To address these gaps, the present study compared adults with misophonia to adults with clinically significant emotion dysregulation across clinical, emotional, and physiological domains using self-report, clinician-rated, and psychophysiological measures collected during an experimental sound paradigm. The primary aim was to characterize similarities and differences between these groups and clarify the extent to which emotional difficulties associated with misophonia resemble those observed in clinical populations with significant emotional dysregulation. Based on prior findings, we hypothesized that both groups would demonstrate elevated levels of distress and impairment. However, we expected the emotion dysregulation group to exhibit greater global emotion dysregulation, psychopathology, and contextual stress burden than the misophonia group. Conversely, we expected the misophonia group to demonstrate substantial impairment despite lower levels of generalized psychopathology and emotional dysfunction. We also examined associations between misophonia severity and emotion sensitivity to evaluate whether misophonia-specific severity was related to broader tendencies toward emotional reactivity.

Finally, we evaluated differences between groups experimentally, through the use of an evocative sound rating task while physiological indices of autonomic functioning were recorded. Prior research demonstrates that individuals with misophonia exhibit heightened autonomic responses to trigger sounds relative to neutral and generally aversive stimuli (Edelstein et al., 2013; Neacsiu et al., 2024). Since extant literature suggests that emotional disturbances in misophonia may differ from broader emotion dysregulation, we anticipated that the misophonia group would exhibit heightened parasympathetic activity (e.g., elevated high frequency heart rate variability) compared to the emotion dysregulation group, reflective of sustained regulation efforts.

## Methods

### Participants

Participants were recruited for an intervention study approved by the [Redacted for Masked Peer Review] University Health System Institutional Review Board, pre-registered under Clinical Trials ID (NCT04348591), and conducted between October 2020 and May 2022. Recruitment was most successful via social media; however, many participants with misophonia also found the study through self-guided internet searches. Participants were paid up to $250 for completing all study components. To determine eligibility after initial online screening and phone screening, participants were evaluated using the Structured Clinical Interview for DSM-5 Disorders (SCID-5; First et al., 2015) and were excluded for active substance use, psychotic disorders, and Bipolar I. Clinicians also rated their overall impairment through a global rating, the Social and Occupational Functioning Assessment Scale (SOFAS; Goldman et al., 1992). For compensation and other screening procedures, see the parent study [Redacted for masked peer review].

Misophonia symptom severity was defined as a score of at least two on each subscale of the Misophonia Questionnaire (MQ; Wu et al., 2014) and a score ≥ 7 on the MQ severity scale. Participants were included in the control group if they scored 89 or higher on the Difficulties in Emotion Regulation Scale (DERS; Gratz & Roemer, 2004) and met criteria for a DSM-5 disorder, consistent with prior work with transdiagnostic emotion dysregulation (Neacsiu et al., 2024). Participants in the misophonia group were not included or excluded based on emotion dysregulation; however, participants in the emotion dysregulation group were excluded if they scored above 6 on the MQ.

### Self-Report Surveys

Participants completed surveys within different domains of misophonia and emotion dysregulation:

1. Participants completed questionnaires to assess for misophonia and hearing related severity: MQ (Wu et al., 2014), Hyperacusis Questionnaire (HQ; Fackrell et al., 2015).
2. To assess for difficulties with emotion dysregulation and clinical impairment, participants completed: DERS (Gratz & Roemer, 2004), the Emotion Regulation Questionnaire (ERQ; Gross & John, 2003), Affect Intensity Measure (AIM; Larsen et al., 1986), Disgust Propensity and Sensitivity Scale-Revised (DPSS-R; Fergus & Valentiner, 2009), Anxiety Sensitivity Index (ASI; Reiss et al., 1986), Inventory of Depression and Anxiety Symptoms-II (IDAS-II; Watson et al., 2012).
3. Participants completed surveys concerning self efficacy and tolerance of negative emotions: PROMIS Self-Efficacy Managing Emotions (PROMIS-SEME; Gruber-Baldini et al., 2017), Distress Tolerance Scale (DTS; Simons & Gaher, 2005).
4. Participants completed surveys concerning the impact of their mental health related symptoms and wellbeing: the Work and Social Adjustment Scale (WSAS; Mundt et al., 2002), the Outcome Questionnaire-45 (OQ-45; Lambert et al., 1996)
5. Finally, participants completed surveys concerning adjustment to a contextual stressor: Coronavirus Impact Scale (CIS; Stoddard et al., 2023), and the Acute Stress Disorder Scale adapted for COVID-19 (ASDS-COVID; Bryant et al., 2000).

### Evocative Sound Challenge Task

Eligible participants completed an Evocative Sound Challenge Task (hereafter referred to as the ‘sound task’) in a sound-attenuated room using noise-canceling headphones. The MATLAB-programmed task included a 3-minute task baseline, a 22-minute sound task, and a 3-minute post-task period. A total of 89 sounds were presented in randomized order, including 28 misophonic, 31 aversive, and 30 neutral stimuli. Sounds were acquired from the International Affective Digitized Sound System (Bradley & Lang, 2007) and other prior literature (Kumar et al., 2017), and each sound was rated by each participant for negativity and misophonic relevance. Participants were presented with a 6-second baseline prior to each sound, a 6-second sound presentation, followed by a rating period (see Figure 1). See the supplement for additional task and stimulus information.

### Psychophysiological Data

Heart rate (HR) and galvanic skin response (GSR) were recorded during the sound task using a BIOPAC MP150 unit (Goleta, CA). AcqKnowledge software was used to collect and pre-process raw electrocardiogram data using a built-in automated heart rate variability (HRV) analysis tool that follows established frequency-domain algorithm guidelines (Camm et al., 1996). High-frequency HRV (HF-HRV), skin conductance level (SCL), and skin conductance responses (SCRs) were extracted for each participant’s baseline, sound task, and recovery period. HF-HRV and SCL values were log10-transformed prior to analysis (see Analysis Plan).

### Analysis Plan

Analyses in this manuscript are secondary to the original paper [Redacted for masked peer review]. In this study, we compared groups on measures of emotion regulation using self-report and psychophysiological assessments. We also compared groups on psychopathology and sensitivity to emotions and chronic stressors, including COVID-19 related stress given the data collection period. Analyses were conducted using IBM SPSS v.31. We conducted independent-samples *t*-tests and chi-square independence tests on self-report measures and applied Bonferroni corrections for multiple comparisons. Outcome measures were assessed for normality using Shapiro-Wilk tests (W < .90) and all passed. HF-HRV and SCL featured significant right skew and were log10-transformed to approximate a normal distribution. We conducted independent samples *t*-tests and ANCOVA on physiological measures, examining whether there were differences in outcomes by group and by phase (baseline, during task, and postline). ANCOVAs were conducted to characterize broad task-phase differences between groups in physiological metrics (HF-HRV and SCL) while controlling for their respective baseline values. Participants with 0 SCRs (n = 19) were removed from SCR analyses as they were considered non-responders.

### Sample

The sample consisted of 60 adults with elevated misophonia impairment (n = 30) or clinically significant emotion dysregulation without misophonia (n = 30). Across groups, there were 5 men, 52 women, and 3 nonbinary adults between the ages of 18 and 54 (M = 28.42; SD = 9.02 years old). The majority of the sample was white. For further demographic detail, see Table 1. An independent samples *t*-test confirmed that the emotion dysregulation control group was lower in misophonia severity than the misophonia group (*t*(58) = −10.82, *p* < .001).

**Table 1.**
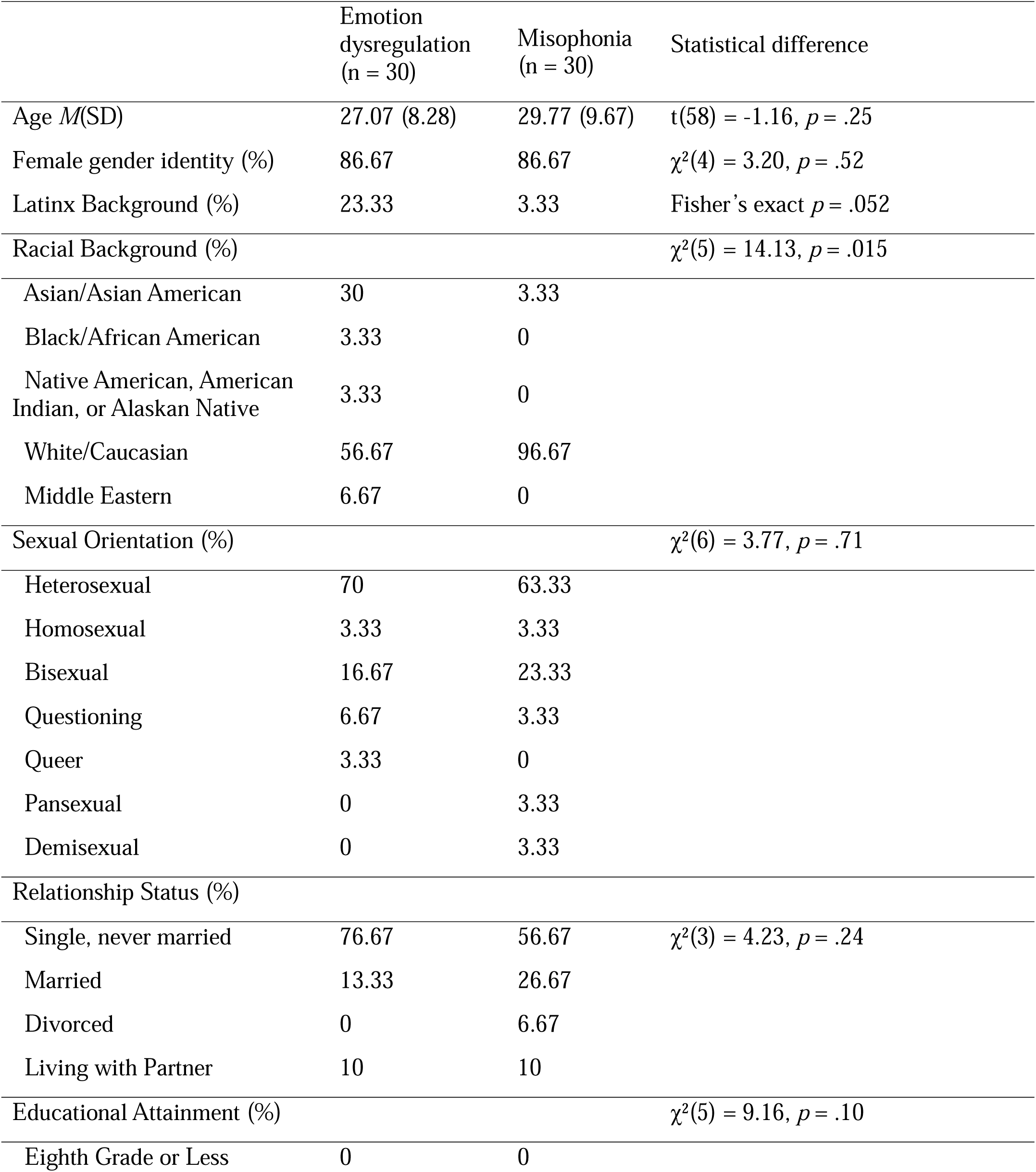
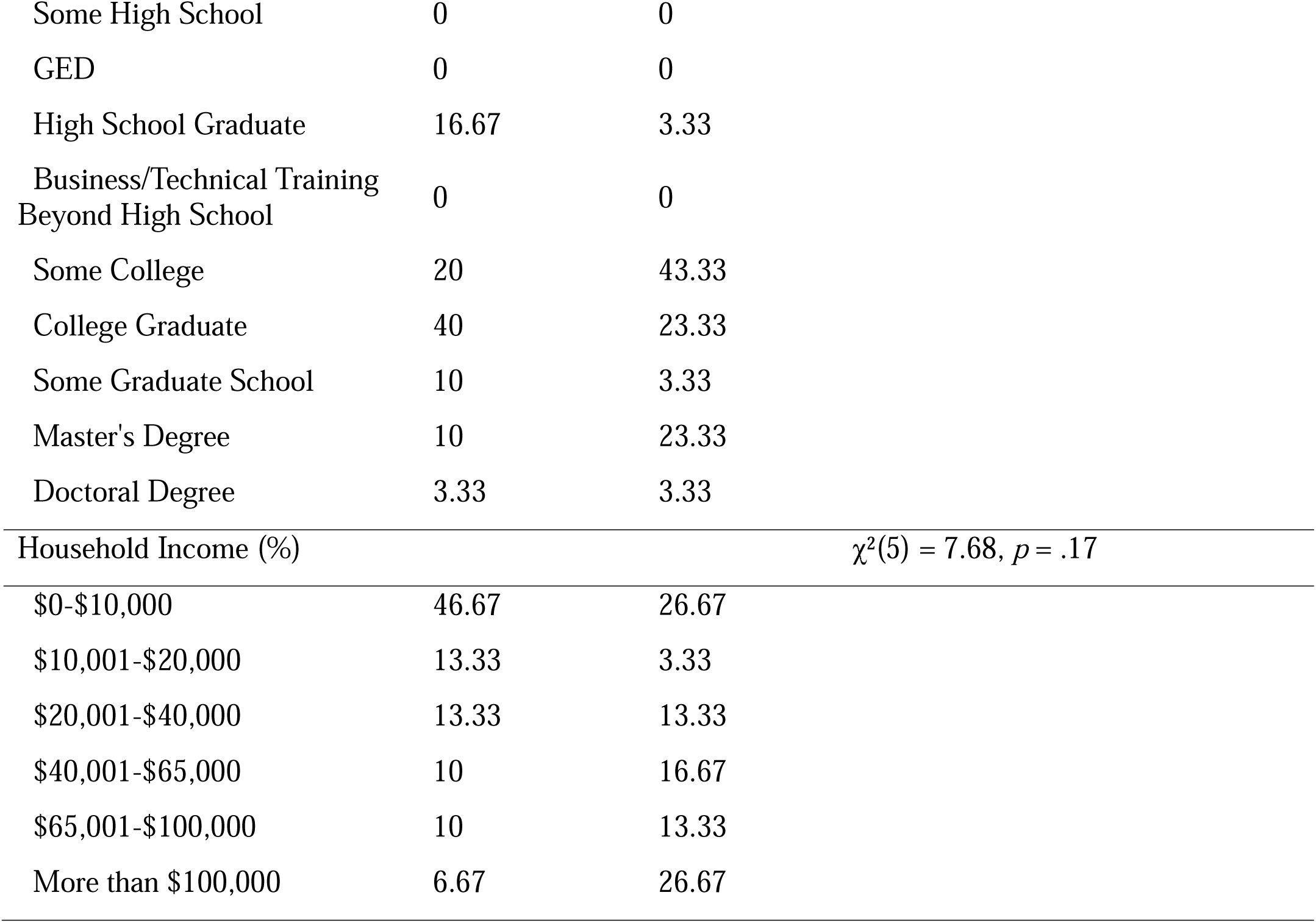
Demographics and Clinical Descriptives by Group.

### Preliminary Analyses

There were no significant differences between groups in age, gender distribution, sexual orientation, marital status, education, income, use of psychotropic medication, current enrollment in psychotherapy, or number of current or lifetime diagnoses met (*p*s >.06; see Tables 1 and 4).

However, the groups differed in racial composition: a higher proportion of participants in the misophonia group identified as White (96.7%) than in the emotion dysregulation group (56.7%; χ²(1, N = 60) = 13.42, *p* < .001) but the proportion of Hispanic/Latinx participants was not statistically different (3.3% vs. 23.3%; Fisher’s exact *p* = .052).

To assess confounding effects of race and ethnicity with imbalanced groups, we created dichotomous race and ethnicity dummy variables (i.e., White/Non-White for racial bins and Hispanic/Non-Hispanic for ethnicity bins). These dichotomized variables were necessary to approximate influence, as the sample was too small and relatively homogeneous to adequately explore all racial and ethnic group differences. Results did not indicate differences between racial or ethnic groups in task phases of HF-HRV, SCL, or SCR (*p*s > .152) and were thus not retained in final analyses for parsimony.

## Results

### Differences in misophonia severity

As a preliminary step, we examined Pearson bivariate correlations among misophonia severity, general emotion dysregulation, and physiological variables. Pearson’s correlations indicated that MQ severity was not significantly associated with HF-HRV, SCLs, SCRs at baseline, task, or postline (*p*s > .133), but it demonstrated a moderate negative correlation with DERS (r(57) = -.361, *p* = .006).

Next, we compared groups on MQ symptom, emotions and behaviors, and severity scores using independent-samples t-tests, and used a Bonferroni correction of .05/3 = .017 to account for multiple comparisons in this group of analyses (Table 2). We found that participants with misophonia reported higher symptom scores (*p* < .001, *d* = 1.75), emotions and behaviors scores, (*p* < .001, *d* = 3.44), and severity ratings (*p* < .001, *d* = 2.79) than clinical controls. All differences remained significant after Bonferroni correction.

**Table 2.** Inclusionary Self-Reports: Misophonia Questionnaire in both samples.

|  | Emotion Dysregulation Group (n=30) *M*(SD) | Misophonia Group (n=30) *M*(SD) | *t*-test |
|---|---|---|---|
| MQ symptom subscale | 1.56 (0.86) | 2.76 (0.46) | *t*(44.14) = −6.77, *p* < .001 |
| MQ emotions and behaviors subscale | 0.89 (0.52) | 2.40 (0.33) | *t*(49.30) = 13.30, *p* <.001 |
| MQ severity | 3.60 (1.77) | 8.63 (1.83) | *t*(58) = -10.82, *p* <.001 |

### Differences in emotion regulation and dysregulation

Next, we compared groups on indices of emotion dysregulation (DERS), emotion regulation (ERQ Reappraisal, Distress Tolerance Scale, PROMIS-SEME), and impairment from emotion dysregulation (WSAS), and used a Bonferroni correction of .05/5 = .01 to account for multiple comparisons in this group of analyses (see Table 3). We found that participants with misophonia reported much higher self-efficacy in emotion regulation than clinical controls (*p* < .001, *d* = 11.16). There were no significant differences between groups after Bonferroni correction in use of reappraisal (*p*_ERQ_Reappraisal_ = .047, *d* = 1.13), functional impairment (*p*_WSAS_ = .078, *d =* 8.62), or access to distress tolerance strategies (*p*_DT_ = .058, *d =* 2.73) (see Table 4).

**Table 3.** Self-Efficacy in Managing Negative Emotions, Reappraisal Ability, Distress Tolerance, and Work and Social Impairment due to Difficulties with Emotion Dysregulation.

| Measure | Emotion Dysregulation Group (n=30) *M*(SD) | Misophonia Group (n=30) *M*(SD) | *t*-test |
|---|---|---|---|
| DERS Total | 112.43 (16.42) | 93.97 (21.42) | *t*(58) = −3.75, *p* < .001 |
| ERQ – Reappraisal | 3.67 (1.04) | 4.27 (1.22) | *t*(58) = −2.03, *p* = .047\* |
| PROMIS–SEME | 68.50 (10.45) | 80.52 (11.83) | *t*(58) = −4.18, *p* < .001 |
| Distress Tolerance Scale | 10.57 (2.29) | 11.93 (3.11) | *t*(58) = −1.94, *p* = .057 |
| WSAS – Emotion Regulation Impairment | 12.43 (8.72) | 8.43 (8.53) | *t*(58) = 1.79, *p* = .078 |

\* = not significant after Bonferroni correction (cutoff = 0.0125)

**Table 4.** DSM-5 disorder count, psychiatric medication, and recent psychotherapy.

| Measure | Emotion Dysregulation Group (n=30) *M*(SD) | Misophonia Group (n=30) *M*(SD) | Difference tests |
|---|---|---|---|
| On psychotropic medications (%) | 40.0 | 36.7 | χ<sup>2</sup>(1) = 0.07, *p* = .791 |
| Recent psychotherapy (%) | 50.0 | 40.0 | χ<sup>2</sup>(1) = 0.61, *p* = .436 |
| Total current diagnoses, *M* (SD) | 2.00 (1.39) | 1.53 (1.80) | *t*(58) = 1.11, *p* = .270 |
| Total lifetime diagnoses, *M* (SD) | 4.14 (2.12) | 3.03 (2.28) | *t*(58) = 1.93, *p* = .059 |

### Physiology During Phases of an Evocative Sound Challenge Task

We also examined between-group differences in emotion regulation using HF-HRV during a baseline resting period, the sound task, and recovery following the sound task. We excluded two participants from these analyses from the misophonia group, one who did not identify any misophonic sounds, and one who did not complete the sound task.

At baseline, misophonic participants had significantly higher HF-HRV than controls (*t*(56) = 2.39, *p* = .010, mean difference = 0.32). The groups also differed in task reactivity, however, the association flipped and the emotion dysregulation group (EMM = 0.81, *SE* = 0.49, 95% CI [0.71, 0.91]) demonstrated higher HF-HRV during task than the misophonia group (EMM = 0.63, *SE* = 0.52, 95% CI [0.52, 0.73]), after controlling for baseline HF-HRV (*F*(1,54) = 6.02, *p* = .017, ηp² = .101). After the sound task, during postline, participants with misophonia (EMM = 1.66, *SE* = 0.06, 95% CI [1.55, 1.78]) exhibited significantly lower recovery HF-HRV than participants with emotion dysregulation (EMM = 1.93, *SE* = 0.06, 95% CI [1.82, 2.05]), *F*(1, 54) = 10.35, *p* = .002, ηp² = .161, when controlling for baseline HF-HRV.

We also examined between-group differences in emotional sensitivity by assessing arousal, as measured by skin conductance level (SCL) and the number of skin conductance responses (SCRs), during rest, the sound task, and recovery following the sound task. The same two participants were excluded from these analyses as mentioned before.

There was insufficient evidence to suggest that SCL differed between groups at baseline. Baseline SCL did not differ significantly between groups, Welch’s *t*(38.96) = -1.02, *p* = .157. Controlling for baseline, participant did not differ in their SCL during the sound task, (*F*(1, 54) = 0.60, *p* = .814, ηp² = .001), or following the sound task (*F*(1, 54) = 3.04, *p* = .087, ηp² = .053).

After removing participants who were non-responders to SCR, there were no differences in SCRs between groups at baseline (*t*_SCRs_(37) = 1.40, *p* = .085; M_SCRs_misophonia_=5.43, *SD* = 3.89; M_SCRS_Emotion dysregulation_ = 3.72, *SD* = 3.66), during the sound task, controlling for baseline (*F*(1, 36) = 0.16, *p* = .691, ηp² = .004), or following the sound task (*F*(1, 33) = 0.03, *p* = .862, ηp² = .001).

### Differences Between Groups in Psychopathology

We examined between-group differences in psychopathology using data from the SCID-5, the OQ-45, and the Hyperacusis Questionnaire, and applied a Bonferroni correction of .05/3 = .017, presented below. See Tables 4-5 for full information.

**Table 5.** Self-reported Outcomes Questionnaire.

|  | Emotion Dysregulation Group (n=30) *M*(SD) | Misophonia Group (n=30) *M*(SD) | *t*-test |
|---|---|---|---|
| OQ – Interpersonal relations | 14.10 (6.38) | 12.13 (6.00) | *t*(58) = - 1.23, *p* = .224 |
| OQ – Social role functioning | 13.00 (4.09) | 10.03 (4.33) | *t*(58) = - 2.73, *p* = .008 |
| OQ-TOTAL | 61.67 (21.54) | 53.16 (21.95) | t(58)= -1.51, *p* = .135 |

### DSM-5 Disorder Count

Across both groups, participants met criteria for an average of 1.76 (SD = 1.61) current diagnoses and 3.58 (SD = 2.25) lifetime diagnoses on the SCID-5. Independent samples *t*-tests indicated no group differences in the number of current diagnoses (*p =* .270) with the misophonia group and emotion dysregulation group showing comparable rates of current diagnoses count. Similarly, rates of psychotropic medication use, and recent psychotherapy did not differ between groups (all *p*s > .44) (See Table 4). There was a difference between groups in the mean clinician SOFAS score *t*(58) = 3.155, *p* = .003, whereby participants in the Misophonia group (M = 78.67, SD = 11.47) had higher scores than participants in the Emotion Dysregulation group (M = 70.36, SD = 8.72) on SOFAS scores. Anxiety disorders were the most common current diagnoses in both groups as demonstrated in the original paper and specifics of disorder breakdowns are listed there [Redacted for masked peer review].

### Functional impairment

We also examined differences in psychopathology using an established self-report measure, the OQ-45. Independent samples *t*-tests indicated that groups did not differ on OQ Total score (*p* = .135), with the misophonia group and emotion dysregulation group showing comparable severity. However, a significant difference emerged for impairment in social role functioning (*p* = .008, *d*= *-*.318), with the misophonia group reporting lower impairment than the emotion dysregulation group. There were no other significant differences found between other subscales (see Table 5).

### Hyperacusis

Participants in the misophonia group reported significantly higher scores on the Hyperacusis Questionnaire (HQ) total score (*M* = 24.17, *SD* = 7.17) compared with participants in the emotion dysregulation group (*M* = 10.33, *SD* = 5.57), *t*(58) = 8.34, *p* < .001, *d* = 2.15.

### Differences Between Groups in Emotion Sensitivity

We examined between-group differences in indices of sensitivity to specific emotions (anger, anxiety, disgust), affect intensity, as well as sensitivity to a global, chronic stressor (distress induced by COVID-19 pandemic) (see Table 6). Because we examined 4 measures under the construct of emotional sensitivity, we used a Bonferroni correction (*p* = .013) to interpret these results. After the Bonferroni correction, we did not find any differences between groups in sensitivity to specific emotions. Participants in the misophonia group reported higher anger sensitivity (*M*_IDAS_ = 8.73, *SD*_IDAS_ = 3.42) and lower anxiety sensitivity (*M*_ASI_ = 17.67, *SD*_ASI_ = 9.84) compared with participants in the emotion dysregulation group (*M*_IDAS_ = 6.97, *SD*_IDAS_ = 2.54; *M*_ASI_ = 23.67, *SD*_ASI_ = 12.17), *t*_IDAS_(58) = 2.27, *p* = .027; *t*_ASI_(58) = −2.10, *p* = .040, however these did not survive Bonferroni correction. Participants did not differ in affect intensity between groups in either total of AIM or the subscales (*p*s = .338 - 0.972).

**Table 6.**
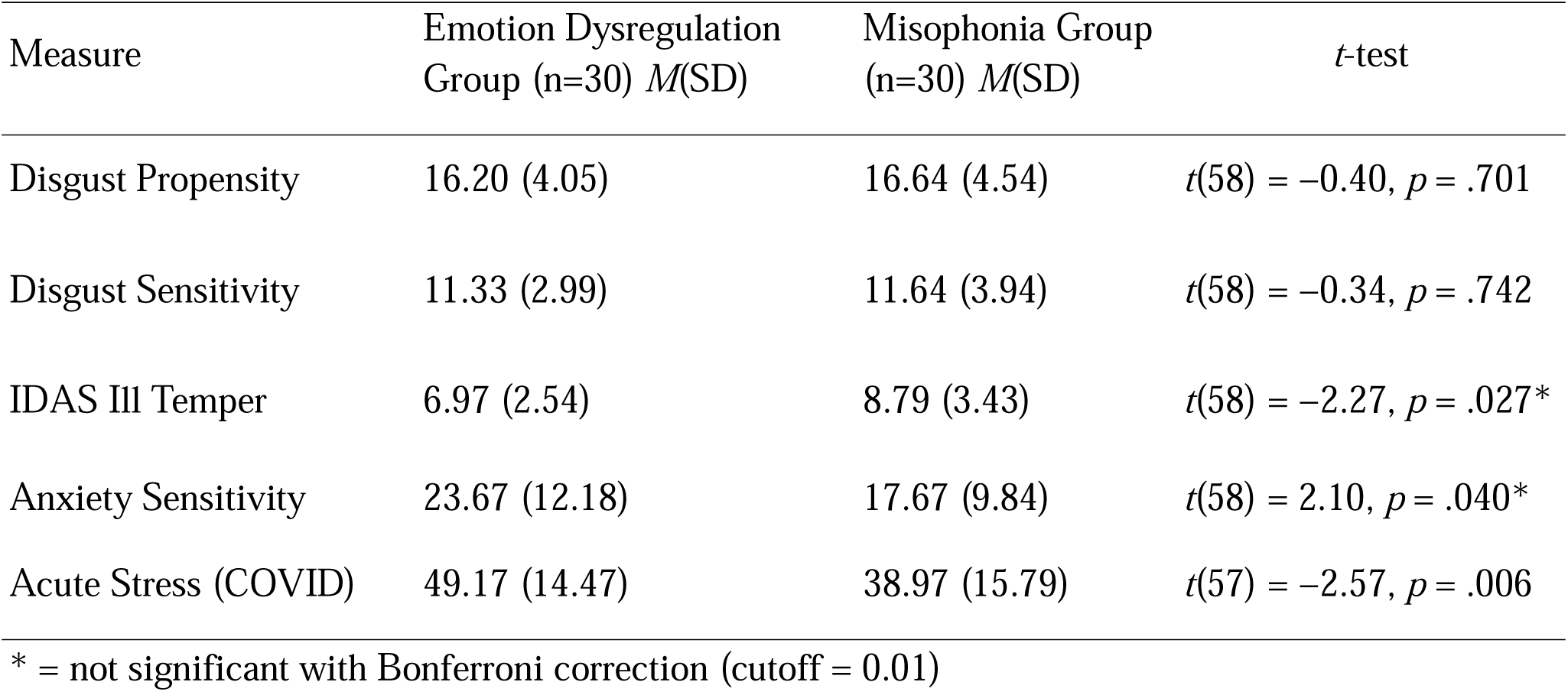
Comparison of Sensitivity to Negative Emotions and Contextual Stress Between Groups.

Participants with misophonia demonstrated significantly lower sensitivity to a chronic stressor when compared to clinical controls. Specifically, participants in the misophonia group reported lower scores on the Acute Stress Disorder Scale adapted for COVID-19 compared with participants in the emotion dysregulation group (*p* = .006). There was no significant group difference in COVID-related impairment severity (*p* = .120). The MQ subscales did not correlate with any emotional sensitivity indices significantly using bivariate correlations (all *p*s > .84).

## Discussion

The aim of the present study was to clarify whether emotional difficulties associated with misophonia resemble broader clinical patterns or a more circumscribed pattern of dysfunction. Although adults with misophonia demonstrated clinically meaningful impairment with substantial psychiatric comorbidity and elevated emotion dysregulation, they reported lower psychopathology, lower global emotion dysregulation, and greater emotion regulation self-efficacy than clinical counterparts. Furthermore, participants with misophonia reported significantly lower sensitivity to a global, chronic stressor (i.e., COVID-19-related stress) than clinical controls. Collectively, these findings suggest that emotion regulation is clinically relevant to misophonia, consistent with prior research (Dixon et al., 2024; Erfanian et al., 2018; Guetta et al., 2022), while suggesting that the overall pattern of emotional functioning may differ from the more pervasive dysfunction characteristic of clinically dysregulated groups.

The pattern of psychiatric symptom severity suggests that misophonia is accompanied by a similar profile of symptomatic breadth, but less perversive functional impairment. For instance, in our sample, anxiety disorders were ubiquitous in both groups. This is similar to phenotyping research demonstrating higher rates of anxiety disorders than would be expected in the general population (Rosenthal et al., 2022) and higher misophonia severity being significantly correlated with anxiety scores, but also depression and obsessive compulsive disorder (Brout et al., 2018). In addition, they did not differ in the number of current diagnoses, in rates of psychotropic medication use or recent psychotherapy, or in self-reported distress and interpersonal impairment. At the same time, our results suggest the groups did diverge in functioning rather than distress burden. While endorsing comparable degrees of general distress on surveys, participants in the Misophonia group were rated as substantially less impaired by clinicians (SOFAS) and participants self-reported less impairment in social role functioning.

This interpretation of specificity is also supported by the results of The Hyperacusis Questionnaire, wherein Misophonia participants scored more than two standard deviations above the emotion dysregulation group (*d* = 2.15). This could indicate that heightened sensitivity to and intolerance of everyday sound is not a general feature of clinical emotion dysregulation. This is similar to prior findings of phenotyping research indicating hyperacusis was significantly correlated with Misophonia (Rosenthal et al., 2022) and in a treatment sample demonstrating considerable overlap between the hyperacusis and misophonia such that those being treated for hyperacusis should be screened for misophonia as well (Aazh et al., 2022). Another study examined if emotion dysregulation predicted misophonia and provided evidence that difficulties with emotion dysregulation may differentiate the two (Aryal et al., 2025). Adults with misophonia carried a comparable diagnostic and symptomatic load to a psychiatrically severe comparison group yet were less globally impaired and were distinguished by sound-specific sensitivity rather than by generalized emotional disturbance. These findings speak to a long-standing interpretive problem in this literature whereby emotional difficulties in misophonia have most often been benchmarked against non-clinical comparison groups. Together, these findings are consistent with the possibility that misophonia is prevalent across different diagnostic groups, and that the difficulties associated with it are not readily accounted for by any one psychiatric disorder or by generalized psychopathology.

Several of our findings converge on the conclusion that misophonia is not characterized by uniformly heightened emotional sensitivity. For instance, adults with misophonia in our sample did not differ from the clinical comparison group in overall affect sensitivity, or sensitivity to anger, anxiety, or disgust, and MQ subscale scores were unrelated to any index of emotional sensitivity. They also reported *lower* distress in response to a prolonged real-world stressor, the COVID-19 pandemic, than adults selected for clinically significant emotion dysregulation. These results align with ecological momentary assessment work showing that adults with misophonia report relatively low negative affect outside of periods of trigger exposure (Shan et al., 2026). Across self-reports, the threshold at which negative emotion is elicited does not appear to be broadly lowered in misophonia, which the electrodermal data support. In our sample, neither skin conductance level nor the frequency of skin conductance responses differed between groups at rest, during the sound challenge, or during recovery. This was a partially surprising finding given prior findings that other research has shown misophonic participants to have elevated skin conductance responses to misophonic cues relative to aversive cues (Edelstein et al., 2013), greater skin conductance and heart rate responses to trigger sounds than controls (Kumar et al., 2017), and more frequent and larger skin conductance responses to trigger sounds than controls (Grossini et al., 2022).

At the same time, findings are not uniform: in a multi-method experimental study, skin conductance only partially confirmed heightened arousal to trigger sounds (Siepsiak et al., 2023), and a recent study found no group differences in skin conductance despite group differences in HRV (Rinaldi et al., 2026). Notably, each of these studies compared individuals with misophonia with non-clinical controls; the absence of electrodermal differences here may partly reflect the use of a clinically dysregulated comparison group. At the same time, the individual components of the sound task were not examined in the present analysis. As a result, it is possible that differences in responding between groups across sound categories were obscured when responses were aggregated at the level of the broader sound task. Such differences may become apparent in more fine-grained analyses examining responses to individual sound categories.

Despite this more circumscribed pattern of emotional reactivity, prior work has consistently documented elevated emotion dysregulation in misophonia (Cassiello-Robbins et al., 2020; Wu et al., 2014). We argue the present findings reinforce its clinical relevance while qualifying its interpretation. Similarly, there could be two different ways to account for this: either in terms of the role of emotion dysregulation in misophonia, or in terms of the interpretation people with misophonia have about emotion dysregulation.

One possibility is that emotion dysregulation constitutes a meaningful problem in misophonia and contributes to the maintenance of misophonic distress beyond the immediate response to trigger exposure. Consistent with this possibility, misophonia has been associated with greater affective instability and difficulties with emotion regulation (Guetta et al., 2022), and difficulties controlling impulsive behavior when distressed partially mediate the association between neuroticism and misophonia (Cassiello-Robbins et al., 2020). In a nationally representative sample, DERS scores were significantly associated with misophonic sensitivity, reactions, and misophonia-related impairment (Dixon et al., 2024). Similarly, it has been shown to account for the association between childhood trauma related impairments in emotion regulation and misophonia (Kılınçel et al., 2026). Moreover, among adults with clinically significant misophonia, severity was specifically associated with difficulties in goal-directed behavior, impulse control, access to regulation strategies, and nonacceptance of emotions (Aryal et al., 2025).Three of these (goal-directed behavior, impulse control, and nonacceptance) are the dimensions on which our misophonia group did not differ from the clinical comparison group.

Alternatively, global self-report measures of emotion dysregulation may partly capture the accumulated burden of trigger-related emotional experiences, even though these measures are intended to index broader regulatory difficulties. Individuals with misophonia may disproportionately anchor judgments about their regulatory ability to situations involving highly distressing triggers, possibly elevating estimates of generalized emotion dysregulation even when regulatory difficulties are less apparent in other contexts. The observation that participants with misophonia reported comparatively low emotion dysregulation alongside impairment that did not differ significantly from that of the clinical comparison group may be more consistent with this second possibility, although the present design does not allow us to distinguish between these interpretations.

Recent biopsychosocial models conceptualize misophonia as arising through interactions among sensory sensitivity, attentional processes, appraisal, and emotion regulation (Hanna & Rosenthal, 2026). Within this framework, regulatory difficulty would be expected to emerge specifically where trigger cues have acquired heightened emotional salience. Misophonic distress may therefore relate more closely to salience attribution, expectancy, attentional allocation, appraisal, and the personal meaning assigned to trigger sounds than to generalized regulatory deficit, an account consistent with models emphasizing aberrant salience and attentional deployment (Kumar et al., 2017; Schröder et al., 2019), as well as with recent ecological momentary assessment work documenting substantial within-person fluctuation in regulatory capacity in misophonia (Pavlacic et al., 2026).

Importantly, greater regulatory demand in response to trigger exposure should not necessarily be equated with deficient regulatory capacity overall. The parasympathetic findings were the one domain in which the groups clearly diverged. At rest, adults with misophonia showed higher HF-HRV than the emotion dysregulation group, consistent with the self-report picture of comparatively preserved general regulatory capacity. Following the sound challenge, however, this pattern reversed: after adjusting for baseline, the misophonia group showed significantly lower HF-HRV both during the sound task and at postline. Higher HRV has generally been associated with regulatory capacity and resilience (Brown et al., 2022; Thayer et al., 2012), and interventions that raise HRV have been linked to reductions in stress and anxiety (Goessl et al., 2017).

This pattern of findings could be accounted for by the sound challenge being more taxing for participants with misophonia. The sound task was specifically disorder-relevant for the misophonia group, consistent with prior evidence of heightened physiological responses to disorder-specific sounds (Oszczapinska et al., 2025), whereas it represented a more general stressor for the emotion dysregulation group. Thus, lower HF-HRV during and after exposure in the misophonia group may reflect greater regulatory demand imposed by personally salient triggers rather than deficient regulatory capacity overall. This interpretation is consistent with the broader pattern of results, in which difficulties in misophonia appeared to be organized around trigger cues rather than distributed across emotional contexts. Alternatively, the findings could reflect vagal withdrawal or less effective parasympathetic re-engagement in misophonia, sustained regulatory efforts in the emotion dysregulation group, or even more efficient (and therefore lower magnitude) regulation to typically triggering stimuli in the misophonia group.

Although misophonia symptom severity was not associated with any physiological indices in the study, group differences in cardiac physiology suggest that the groups may have responded differently to the sound challenge. These accounts are difficult to separate in the present design. Participants were not instructed to use any particular emotion regulation strategy, so physiological responses cannot be interpreted as direct indicators of successful or unsuccessful deliberate regulation attempts. Moreover, the groups were not exposed to stimuli of comparable personal relevance.

The electrodermal data further constrain this interpretation. Because SCL and SCR did not differ between groups at any point, the HF-HRV differences occurred within a context of comparable measured sympathetic arousal. As noted above, Rinaldi et al. (2026) similarly found lower HRV in individuals with misophonia without corresponding differences in skin conductance, although in that study the HRV differences were present across both sound and silence conditions rather than being tied to trigger exposure. Taken together, subjective experience and autonomic responding did not covary uniformly, which argues against a simple generalized hyperarousal account of misophonia. Still, the present data cannot fully establish whether the HF-HRV differences reflect less effective regulation, greater regulatory demand imposed by disorder-relevant stimuli, or other context-dependent processes, a question that will require designs in which regulatory strategy is instructed and stimulus relevance is matched across groups.

### Limitations

Several additional limitations should be considered when interpreting the present findings. First, both groups were predominantly composed of white women, reflecting a pattern commonly observed in the misophonia literature. Consequently, the generalizability of these findings to more diverse populations remains limited. Second, although the present findings support distinctions between generalized emotion dysregulation and trigger-specific responding, the physiological analyses were not designed to characterize moment-to-moment regulatory trajectories. Future studies using repeated measurements, longer recovery windows, and stimulus-specific modeling may more precisely characterize activation and recovery dynamics, including whether regulatory processes differ across trigger types or interpersonal contexts.

Third, although the misophonia group demonstrated lower overall emotion dysregulation than the clinical comparison group, a subset also met criteria for clinically significant emotion dysregulation. The study was not sufficiently powered to examine this subgroup separately; larger samples are needed to determine whether individuals with both misophonia and clinically significant emotion dysregulation differ from those with misophonia alone. Similarly, the cross-sectional design precludes determining whether lower psychopathology and emotion dysregulation in the misophonia group reflect qualitative differences between conditions or different stages in the development of broader dysregulation. Longitudinal studies are needed to clarify whether generalized emotion dysregulation precedes, follows, or develops alongside chronic misophonic distress. Finally, global self-report measures may disproportionately capture participants’ trigger-related experiences rather than their broader emotional functioning, potentially inflating estimates of generalized emotion dysregulation. Integrating self-report with behavioral and physiological measures may therefore help distinguish generalized dysregulation from context-specific regulatory difficulties.

## Conclusion

By comparing adults with misophonia with a clinically dysregulated comparison group, the present study provides a more stringent test of whether the emotional difficulties associated with misophonia resemble generalized emotion dysregulation. Despite clinically meaningful emotion dysregulation and substantial psychiatric comorbidity, adults with misophonia reported lower global emotion dysregulation and greater regulation self-efficacy and were rated as less functionally impaired yet did not differ from the comparison group in number of current diagnoses, self-reported symptom distress, sensitivity to specific emotions, or electrodermal responding. Notably, impairment attributable to emotional difficulties did not differ significantly between groups, and adults with misophonia showed lower baseline-adjusted HF-HRV following exposure to personally relevant sounds. Together, these findings do not suggest that misophonia is characterized by uniformly heightened emotional sensitivity or uniformly poorer emotional regulation. Group differences instead appeared to depend on the salience of disorder-relevant cues, raising the possibility that context-general measures of emotion regulation may not adequately capture the difficulties that matter most in misophonia. Emotion regulation therefore appears to be an important component of misophonia, but clarifying its role will require attention to how regulatory processes interact with attention, appraisal, and the personal meaning assigned to trigger sounds.

## Data Availability Statement

The data that support the findings of this study are available from the corresponding author upon reasonable request.

## Authorship Contributions

ZEB: Conceptualization, Formal Analysis, Investigation, Validation, Visualization, Writing-original draft, Writing-review & editing. JYC: Conceptualization, Data Curation, Data Processing, Investigation, Validation, Writing-editing & review. SCN: Formal Analysis, Investigation, Supervision, Writing-editing & review, Writing-original draft. ADN: Acquisition of Funding, Conceptualization, Data Curation, Investigation, Methodology, Project Administration, Resources, Software, Supervision, Writing-original draft, Writing-review & editing.

## Data Availability

All data produced in the present study are available upon reasonable request to the authors

## Acknowledgments

Data from the present paper were also presented in several conference talks. This research and the completion of the manuscript were supported by a Misophonia award granted to the last author by the REAM Foundation. The authors would like to thank the participants who took part in this study and to acknowledge Lisalynn Kelley, Judit Wright, Eleanor Wood, John Powers, PhD, John Graner, PhD, Simon Davis, PhD, Sukhbinder Kumar, PhD, Deepika Anand, PhD, Brenden Li, Victoria Szymkiewicz, Joshua Kang, and our research assistants and the DUMC CMER, BIAC, and BSRC staff for their contributions.

