## Supplemental Information for "Comparing Misophonia with Clinical Emotional Dysregulation: A Multi-method Characterization Study"

### *Demographics*

An independent-samples *t*-test provided insufficient evidence to suggest that age differed significantly between the misophonia group ($M = 29.77$, $SD = 9.67$) and the emotion dysregulation group ($M = 27.07$, $SD = 8.28$), $t(58) = 1.16$, $p = .250$, $d = 0.30$.

There was a significant association between racial background and group membership, $\chi^2(1, N = 60) = 17.33$, $p < .001$, such that the misophonia group included a higher proportion of White participants (93.3%) compared to the emotion dysregulation group (43.3%). For Hispanic/Latinx ethnicity, the chi-square test indicated a significant association between groups, $\chi^2(1, N = 60) = 5.19$, $p = .023$; however, because of small cell counts, Fisher's exact test was conducted and was not statistically significant (two-sided $p = .052$), the misophonia group consisted almost entirely of non-Hispanic/Latinx participants (96.7%), compared to 76.7% in the emotion dysregulation group.

### *Misophonia Presentation and Severity*

At intake, 27 participants identified themselves as having difficulties with misophonia. These participants reported an average onset for misophonic distress at 12.22 years of age (Range: 7-30; SD: 6.06). Fewer than half (36.7%) reported sound sensitivity to their primary care provider, and only 40% sought help with this problem. Two additional participants did not self-identify as misophonic but scored above cutoff on the MQ, rendering the final sample size to 29.

### *Evocative Sound Challenge Task*

The task was programmed in MATLAB and completed in a sound-attenuated room through noise-canceling headphones. It consisted of a 3-min resting baseline, the 22-min sound task, and a 3-min post-task recovery period (see Figure 1). During the sound task, 89 sounds (28 misophonic, 31 aversive, and 30 neutral) were presented in randomized order. Each trial began with a 6-s pre-sound baseline, followed by a 6-s sound presentation (sounds were trimmed or looped to a 6-s duration) and a rating period in which participants rated each sound for negativity and misophonic relevance.

Sounds were drawn from the International Affective Digitized Sounds (IADS; Bradley & Lang, 2007), a free online sound database (fesliyanstudios.com), and sounds described as misophonic triggers in the published literature (Kumar et al., 2017). Aversive and neutral sounds were selected using IADS normative valence and arousal ratings, and misophonic sounds were selected to match trigger sounds described in prior work.

### *Physiology*

Electrocardiogram (ECG) and galvanic skin response (GSR) were recorded continuously throughout the session with a BIOPAC MP150 system (BIOPAC Systems, Inc., Goleta, CA), using GSR electrodes on the index and ring fingers and ECG electrodes on the wrist and ankle. Signals were sampled at 1 kHz.

*HF-HRV Processing*

High-frequency heart rate variability (HF-HRV) was derived separately for each baseline period, during the Evocative Sound Challenge Task, and post-task period. Because reliable frequency-domain HRV estimation requires recordings of at least approximately 1 minute for the HF component (Camm et al., 1996), HF-HRV could not be calculated for individual 6-s sound presentations. Accordingly, mean HF-HRV for each period was used as the outcome measure.

High-frequency heart rate variability (HF-HRV) was defined as spectral power in the high-frequency band (0.15–0.40 Hz; Camm et al., 1996) and calculated using the single-epoch spectral HRV analysis function in AcqKnowledge. ECG recordings were cleaned prior to analysis, and the resulting tachograms were visually inspected for artifacts. When artifacts were identified, additional ECG cleaning was performed before repeating the analysis. HF-HRV was extracted for the baseline, 22-minute sound task (including rating periods), and post-task periods. HF-HRV values were log10-transformed prior to analysis.

*SCL and SCR Processing*

Skin conductance data were processed using AcqKnowledge and MATLAB. The GSR signal was low-pass filtered at 1 Hz using a finite impulse response (FIR) filter with a Blackman window, then resampled to 10 samples/s using linear interpolation. A custom MATLAB script was used to isolate tonic skin conductance level (SCL) by tracking local minima and removing phasic responses. The resulting SCL signal was visually inspected and reimported into AcqKnowledge. Mean SCL was extracted for each task period.

Skin conductance responses (SCRs) were identified as increases in conductance of at least 0.02 μS, measured from the preceding trough to the response peak. All qualifying SCRs were counted within the 3-minute baseline, 22-minute sound task, and 3-minute post-task periods. Participants with no SCRs across the entire intake task were classified as non-responders and excluded from SCR analyses.

***Clinician Rated Items***

*Structured Clinical Interview for DSM Disorders-5 (SCID).* The SCID is organized into diagnostic modules according to the DSM-5 to assess mood, psychotic, substance use, anxiety, obsessive-compulsive, eating, externalizing, gambling, attention deficit hyperactivity, and trauma- and stressor related disorders (First et al., 2015). The SCID-PD is used to evaluate personality disorders according to the DSM-5 criteria.

*Social and Occupational Functioning Assessment Scale (SOFAS).* The SOFAS scale is a global assessment of overall functioning of a participant completed typically at the end of a structured clinical interview, such as the SCID-5 (First et al., 2015). It is a single numerical item on a scale of 0-100, wherein higher values indicate higher functioning. Scores are grouped into

10-point ranges, from superior functioning (91-100), good (81-90), slight impairment (71-80), some difficulty (61-70), moderate difficulty (51-60), serious impairment (41-50), major impairment (31-40), unable to function (21-30), unable to function independently occasionally (11-20), to unable to function independently persistently (1-10).

### *Self-Report Scales*

*Misophonia Questionnaire (MQ).*The MQ assesses sensitivity to misophonic sounds and the accompanying symptoms in three parts (Wu et al., 2014). The first scale is the Symptom scale, containing 8-items, in which the sensitivities to seven common misophonic trigger sounds are evaluated on a Likert scale from 0 ("Not at all true") to 4 ("Always true"). There is one item in "other" category. The second scale, Emotions and Behaviors Scale, is completed if participants endorse at least "rarely true" (≥ 1) on any item. This assesses frequency and reactions as a result of sound exposure of ten specific reactions and one "other" reaction, using a 5-point scale from 0 ("never") to 4 ("always"). Third, there is a single item for rating the severity of sound sensitivity on a 15-point scale, where higher scores indicate greater severity. Scores 1 to 3 suggest minimum to mild symptoms, whereas scores 13 to 15 suggest very severe symptoms. Internal consistency of the measure with our sample was considered good with Cronbach's α = 0.83.

*Hyperacusis Questionnaire (HQ).* The HQ (Fackrell et al., 2015; Khalfa, 2002) contains two components to measure increased sensitivity to sound. The first consists of questions concerning auditory disorders and noise exposure in binary questions. The second part is comprised of 14 negatively worded items rated on a 4-point Likert scale ranging from 0 ("No") to 3 ("Yes, a lot"). The total score can range from 0-42 and scores higher than 28 indicate hyperacusis. There was excellent internal consistency for this measure in our sample Cronbach's α = 0.90.

*Difficulties with Emotion Regulation Scale.* The DERS (Gratz & Roemer, 2004) contains 36 items to evaluate the frequency of clinically significant emotion regulation challenges on a Likert scale ranging from 1 ("Almost never") to 5 ("Almost always"). Higher scores suggest higher difficulty with emotion regulation. There are six subscales, including nonacceptance of emotions, emotional awareness, access to emotion regulation strategies, goal-directed behavior during emotional arousal, impulse control, and emotional clarity. Within our sample, total scale internal consistency was considered good (α = 0.84), and all subscales had good to excellent internal consistency: nonacceptance of emotions α = 0.90, emotional awareness α = 0.82, access to emotion regulation strategies α = 0.87, goal-directed behavior during emotional arousal α = 0.89, impulse control α = 0.87, and emotional clarity α = 0.90.

*The Emotion Regulation Questionnaire (ERQ).* The ERQ (Gross & John, 2003) is a 10-item scale used to measure management and regulation of emotions on a Likert scale from 1 ("Strongly disagree") to 7 ("Strongly agree"). The measure contains subscales cognitive reappraisal and expressive suppression which are scored separately. Internal consistency with our sample for subscales, ERQ reappraisal (α = 0.89) and ERQ suppression, were good (α = 0.83).

*The Work and Social Adjustment Scale (WSAS).* The WSAS (Mundt et al., 2002) is a 5-Item Likert scale containing questions about the impact of mental health concerns on work, social, relationship, and leisure, rated from 0 ("Not at all") to 8 ("Very severe"). Total scores are calculated as the sum of items. Scores 0-9 indicate low impairment, scores 10-19 indicate moderate impairment, scores 20-40 indicate severe impairment. Internal consistency with our sample was considered excellent (α = 0.90).

*The Outcome Questionnaire-45 (OQ-45).* The OQ-45 is a 45-item battery assessing three core symptom dimensions of life functioning which consist of symptom distress, interpersonal relations, and social role performance in the last week (Lambert et al., 1996). It uses a 5-point Likert scale ranging 0 ("Never") to 4 ("Always"). The total score can range from 0-180 and higher scores indicate greater distress and functional impairment. The sample demonstrated good internal consistency (α = 0.81).

*The Affect Intensity Measure (AIM).* The AIM (Bryant et al., 1996; Larsen et al., 1986) is a 40-item scale evaluating magnitude of emotional responses with a three factor structure comprising of positive affectivity, negative intensity, and negative reactivity. Items are responded to using a 6-point Likert scale ranging from 1 ("Never") to 6 ("Always"). The sample demonstrated good internal consistency (α = 0.88).

*The PROMIS Self Efficacy Managing Emotions (PROMIS-SEME).* The PROMIS-SEME, (Gruber-Baldini et al., 2017) measures confidence in managing emotions, stress, and negative feelings. It contains 25 items and is rated on a 5-point Likert scale from 1 ("I am not confident at all") to 5 ("I am very confident"). Higher scores indicate greater self-efficacy. The sample demonstrated excellent internal consistency (α = 0.90).

*Distress Tolerance Scale (DTS).* The DTS (Simons & Gaher, 2005) measures perceived ability to withstand negative affect or aversive psychological states. It is a 15-item scale rated on a 5-point Likert scale from 1 ("Strongly agree") to 5 ("Strongly disagree"). It is broken into four subscales that include Tolerance, Absorption, Appraisal, and Regulation. Higher total scores indicate higher distress tolerance. The sample demonstrated good internal consistency (α = 0.89).

*Disgust Propensity and Sensitivity Scale-Revised (DPSS-R).* The DPSS-R (Fergus & Valentiner, 2009) consists of 12 total items to which participants respond on a 5-point Likert scale ranging from 1 ("Never") to 5 ("Always") regarding tendency to experience disgust and distress caused by disgust. The scale is broken into two sub scales of Disgust Propensity (DP) and Disgust Sensitivity (DS), with six items per scale. Within this sample, internal consistency was considered good for the total scale (α = 0.85). Internal consistency for subscales Disgust Propensity and Disgust Sensitivity were considered good as well ($\alpha_{\text{Disgust Propensity}} = 0.71$, $\alpha_{\text{Disgust Sensitivity}} = 0.72$).

*Inventory of Depression and Anxiety Symptoms* (IDAS-II) The IDAS-II (Watson et al., 2012)is a broad measurement of internalizing disorders consisting of 64 items assessing components of fear and distress. There are 11 non-overlapping subscales which are dysphoria, insomnia, lassitude/fatigue, suicidality, appetite loss, appetite gain, ill temper (anger/hostility), wellbeing, panic, social anxiety, and traumatic intrusions. Higher scores indicate greater presence of corresponding scale. Participants respond to each item on a 5-point Likert scale from 1 (“Not at all”) to 5 (“Extremely”). In this study we selected the five items used for the anger/hostility subscale (IDAS25, IDAS28, IDAS30, IDAS31 and IDAS33 were used). Within this sample, internal consistency for the anger/hostility subscale was good (α = 0.83).

*Anxiety Sensitivity Index (ASI).* The ASI (Reiss et al., 1986)contains 16 items measuring anxiety related bodily and mental sensations to which participants respond on a Likert scale ranging from 0 (“Very little”) to 4 (“Very much”). Item responses are summed to produce a total score (range 0–64), with higher scores indicating greater anxiety sensitivity. Internal consistency in our sample was considered good (α = 0.88).

*Coronavirus Impact Scale (CIS).* The CIS (Stoddard et al., 2023)is a 12-item self-report measure of how much the COVID-19 pandemic has changed respondents’ lives. Items 1–8 assess change in routines, family income/employment, food access, medical health care access, mental health treatment access, access to extended family and non-family social supports, experiences of pandemic-related stress, and stress and discord in the family. Each is rated on a 4-point scale from 0 (“No change” or “None”) to 3 (“Severe”), with behaviorally anchored descriptions at each level, and summed to form an impact score. Item 9 rates the severity of a personal coronavirus diagnosis (0–3). Items 10 and 11 record the number of immediate and extended family members or close friends diagnosed and rate the symptom severity of the most ill person (1 = managed at home to 4 = died). Item 12 is an open-ended response. Internal consistency for items 1-9 in our sample was acceptable (α = 0.72).

*Acute Stress Disorder Scale adapted for COVID-19.* The ASDS for COVID-19 (Bryant et al., 2000) was adapted to the COVID-19 pandemic, and the ASDS-19 created by the Center for Pediatric Traumatic Stress in March 2020 at the Children’s Hospital of Philadelphia. Items consist of different symptoms of emotional distress and participants are asked to consider how they have been feeling related to COVID-19. It consists of a 20-item inventory rated on a 5-point Likert scale ranging from 1 (“Not at all”) to 5 (“Very much”), where higher scores indicate higher distress. Internal consistency was excellent for this measure (α = 0.93).
